# Biallelic variants in *COX18* cause a mitochondrial disorder primarily manifesting as peripheral neuropathy

**DOI:** 10.1101/2024.07.03.24309787

**Authors:** Camila Armirola-Ricaurte, Laura Morant, Isabelle Adant, Sherifa Ahmed Hamed, Menelaos Pipis, Stephanie Efthymiou, Silvia Amor-Barris, Derek Atkinson, Liedewei Van de Vondel, Aleksandra Tomic, Els de Vriendt, Stephan Zuchner, Bart Ghesquiere, Michael Hanna, Henry Houlden, Michael P Lunn, Mary M Reilly, Vedrana Milic Rasic, Albena Jordanova

## Abstract

Defects in mitochondrial dynamics are a common cause of Charcot-Marie-Tooth disease (CMT), while primary deficiencies in the mitochondrial respiratory chain (MRC) are rare and atypical for this etiology. This study aims to report *COX18* as a novel CMT-causing gene. This gene encodes an assembly factor of mitochondrial Complex IV (CIV) that translocates the C-terminal tail of MTCO2 across the mitochondrial inner membrane.

Exome sequencing was performed in four affected individuals. The patients and available family members underwent thorough neurological and electrophysiological assessment. The impact of one of the identified variants on splicing, protein levels, and mitochondrial bioenergetics was investigated in patient-derived lymphoblasts. The functionality of the mutant protein was assessed using a Proteinase K protection assay and immunoblotting. Neuronal relevance of COX18 was assessed in a *Drosophila melanogaster* knockdown model.

Exome sequencing coupled with homozygosity mapping revealed a homozygous splice variant c.435-6A>G in *COX18* in two siblings with early-onset progressive axonal sensory-motor peripheral neuropathy. By querying external databases, we identified two additional families with rare deleterious biallelic variants in *COX18*. All affected individuals presented with axonal CMT and some patients also exhibited central nervous system symptoms, such as dystonia and spasticity. Functional characterization of the c.435-6A>G variant demonstrated that it leads to the expression of an alternative transcript that lacks exon 2, resulting in a stable but defective COX18 isoform. The mutant protein impairs CIV assembly and activity, leading to a reduction in mitochondrial membrane potential. Downregulation of the *COX18* homolog in *Drosophila melanogaster* displayed signs of neurodegeneration, including locomotor deficit and progressive axonal degeneration of sensory neurons.

Our study presents genetic and functional evidence that supports *COX18* as a newly identified gene candidate for autosomal recessive axonal CMT with or without central nervous system involvement. These findings emphasize the significance of peripheral neuropathy within the spectrum of primary mitochondrial disorders and the role of mitochondrial CIV in the development of CMT. Our research has important implications for the diagnostic workup of CMT patients.

## Introduction

Charcot-Marie-Tooth (CMT) disease is an umbrella term for a group of clinically and genetically heterogeneous sensory-motor peripheral neuropathies. Together, they are the most common inherited neuromuscular disorder, with a prevalence of 9,7-82,3 patients per 100,000 individuals.^1^ Mutations in more than 100 genes, involving all patterns of inheritance, have been reported to cause CMT.^2^ Some of these genes encode proteins that participate in essential nerve-specific processes, such as axonal transport, myelination, and synaptic transmission. Others are however involved in basic cellular pathways (e.g., endosomal trafficking, cell signaling, mRNA processing), and as such, are house-keeping and ubiquitously expressed. It remains unclear why defects in such omnipresent proteins cause a disease predominantly affecting the peripheral nerves.

Alterations in the fission, fusion, and transport of mitochondria are a widely recognized cause of CMT.^3^ In fact, variants in MFN2, a protein involved in mitochondrial fusion, are among the most prevalent causes of axonal CMT (CMT2), accounting for 21-30% of the genetically diagnosed individuals.^4,5^ Variants in *GDAP1*, encoding a protein implicated in mitochondrial fission, are another common cause of CMT2, specifically prevalent in certain geographic regions of the world.^6–9^ Due to their significance, exhaustive clinical and molecular studies examined the natural history and molecular pathogenesis of these CMT2 sub-types.^10–16^ Overall, the general hypothesis is that defects in mitochondrial dynamics impair mitochondrial trafficking, distribution, and turnover along the axons, ultimately leading to bioenergetic failure.^17,18^ Similarly, mitochondrial dysfunction is also a feature of other CMT2 types caused by defects in axonal transport.^19^

In contrast, primary defects in the respiratory chain directly affecting mitochondrial bioenergetics are a rare and relatively unexplored etiology of CMT. For example, a pathogenic variant in the mitochondrial-encoded MT-ATP6, a subunit of ATP synthase (Complex V), has been estimated to account for approximately 1% of unsolved CMT2 patients.^20^ Here, we report biallelic variants in *COX18*, cytochrome c oxidase assembly factor 18, as a novel cause of CMT2. The patients carrying deleterious variants in this gene exhibit progressive sensory-motor polyneuropathy of variable onset and some of them also display signs of central nervous system (CNS) involvement. Functional studies in patient-derived lymphoblasts suggest that the underlying mechanism is a partial loss of function of COX18 that leads to reduced assembly and activity of cytochrome c oxidase (Complex IV, CIV). Downregulation of the COX18’s ortholog in *Drosophila* induces behavioral and neuropathological phenotypes common to other fly models of CMT disease.

## Materials and methods

### Participants

Patients underwent routine neurologic and electrophysiological examinations. This study was approved by the local institutional review boards. All patients signed an informed consent form prior to enrolment.

### Exome sequencing and homozygosity mapping

Genomic DNA was isolated from peripheral blood mononuclear cells according to standard protocols. Exome sequencing (ES) was performed in the two affected members from family 1 using SeqCap EZ Exome Probes v3.0 kit (Roche Holding AG, Basel, Switzerland) for capture. Then, 150 bp exome paired-end sequencing was run on NextSeq 150 platform (Illumina, San Diego, CA). Primary analyses were carried out using the GenomeComb pipeline (version 0.98.3)^21^ which used Burrows-Wheeler algorithm (version 0.7.15-r1140)^22^ for read alignment to hg19 reference human genome and genomic analysis toolkit (GATK) (version 2.4-9-g532efad)^23^ for variant calling. Homozygosity mapping based on WES data was performed as described^24^, using the HOMWES tool from the GenomeComb package (version 0.11.0).^21^ Within the resulting regions of homozygosity, variant filtering and prioritization were performed using GenomeComb^21^ based on a recessive model of inheritance with the following criteria: non-synonymous or splice variants with minor allele frequency (MAF) below 5% and no homozygotes in the control population database gnomAD v2.1.1.^25^ Prioritized variants were evaluated further using Alamut Visual Plus (Sophia Genetics, Switzerland).

### Variant segregation and cohort screening

The resulting variants were confirmed and segregated in the available family members by Sanger sequencing as previously described.^24^ A cohort of 272 unsolved recessive and 90 sporadic CMT patients was screened for deleterious variants in *COX18* using an amplicon target amplification assay (Agilent, https://www.agilent.com). Re-sequencing was performed on a MiSeq (Illumina) platform using 250 bp pair-end reads targeting all exons and exonintron boundaries of the canonical *COX18* transcripts. The primers used are listed in the Supplementary Table 1. All additional variants identified in the cohort screening were validated by Sanger sequencing. Furthermore, GENESIS^26^ and RD-Connect Genome-Phenome Analysis Platform (GPAP) (https://platform.rd-connect.eu/)^27^ were queried online to find additional unrelated CMT patients with *COX18* candidate variants.

### Cell culture

Peripheral blood mononuclear cells from the patients and parents of family 1 were isolated and transformed with Epstein-Barr virus (EBV) as described.^28^ Patient-derived lymphoblasts were maintained as previously described.^29^ Briefly, cell lines were cultured at 37°C, 5% CO_2_ in RPMI 1640 medium containing 15% fetal calf serum, 1% sodium pyruvate, 1% 200M LLglutamine, and 2% penicillin/streptomycin. The cells were passaged every 5 days.

### RNA extraction and cDNA synthesis

Total RNA was extracted from the lymphoblasts using the Universal RNA Purification kit (Roboklon, Berlin, Germany), followed by DNAse treatment with TURBO DNA-free kit (Invitrogen, Thermo Fisher Scientific, Waltham, MA, USA). cDNA was synthesized using iScript Advanced cDNA synthesis kit for RT-qPCR (Bio-Rad, Hercules, CA, USA). *COX18* cDNA was amplified using the primers listed in the Supplementary Table 1.

### Targeted cDNA long read sequencing

Targeted long-read sequencing (T-LRS) of *COX18* cDNA was performed on a MinION platform using a flongle adapter (Oxford Nanopore technologies - ONT, Oxford, UK) as described.^30^ For this purpose, a primer pair was designed to anneal on exon 1 and exon 4, flanking all previously known splicing events of *COX18*. As *COX18* is expressed at low levels in most tissues, PCR amplification with 35 cycles was performed. Sequencing reads were aligned to human genome assembly 38 with minimap2 v5.0.11 in spliced alignment mode. Read splice junction correction, high-confidence isoform definition, and quantification were performed using FLAIR v1.5.^31^

### Mitochondrial isolation

Lymphoblasts were collected from one T175 flask per individual, washed with PBS and resuspended in mitochondrial isolation buffer (250mM mannitol, 0.5mM EGTA, 5 mM HEPES/KOH, pH 7.4). The cells were lysed using 10 strokes of a 1 ml syringe attached to a 26.5G needle. The resulting lysate was centrifuged twice at 600 g for 10 minutes at 4°C. The supernatant was then centrifuged twice at 7000 g for 10 min at 4°C to separate the cytosolic (supernatant) and the mitochondrial fraction (pellet). The mitochondrial pellet was resuspended and centrifuged twice at 10,000 g for 10 min at 4°C.

### Proteinase K degradation assay

Isolated mitochondria were divided equally over four tubes; two were resuspended in regular mitochondrial isolation buffer and the other two with osmotic swelling buffer without mannitol (0.5mM EGTA, 5 mM HEPES/KOH, pH 7.4) to generate mitoplasts. One tube from each condition was treated with 5μg/ml of Proteinase K (Thermo Fisher Scientific) for 20 min at 4°C in a head-over-head rotor. Proteinase K digestion was inhibited with the incubation with 1mM phenylmethylsulfonyl fluoride solution (PMSF, sc-482875, Santa Cruz Biotechnology) on ice for 10 min. The mitochondria and mitoplasts were isolated by centrifugation at 10,000 g for 10 min at 4°C. The resulting pellets were resuspended with Laemmli sample buffer and boiled for 5 min at 95°C. Then, the samples were immunoblotted as explained below.

### Complex IV enzymatic assay

Complex IV activity was measured in mitochondrial fractions as previously described.^32^ Lymphoblasts pellets were obtained from one T175 flask and resuspended in 1ml ice-cold Mega Fb buffer (250 mM sucrose, 2 mM HEPES, 0.1 mM EGTA, pH 7.4) supplemented with 0.08 mM digitonin. Then, the cells were disrupted on an ice slurry with 20 strokes of a Teflon-glass Wheaton homogenizer driven by a Glas-Col High Speed Homogenizer variable speed bench top drill at 1800 rpm. Mitochondrial fractions were isolated by differential centrifugation as described. The resulting pellet was resuspended in ice-cold hypotonic buffer (25 mM potassium phosphate, pH 7.2, 5 mM MgCl_2_) and subjected to three freeze-thaw cycles in dry ice and ethanol slurry. CIV and citrate synthase (CS) activity was measured in a Cary 300 UV-Vis spectrophotometer (model number G9823A) as previously described.^33^ Before measurements, reaction cuvettes with 50mM KPi (pH 7.4) were equilibrated to 30°C. Each enriched mitochondria sample was added, followed by the addition of reduced cytochrome c to a concentration of 15mM. After briefly mixing, the absorbance was immediately measured. Then, K_3_Fe(CN)_6_ was added to 1mM to achieve complete oxidation of cytochrome c, and a final reading was taken. The results were calculated as described.^33^

### Measurement of mitochondrial membrane potential

Lymphoblasts were washed twice with prewarmed phosphate-buffered saline (PBS) and incubated with 20uM TMRE (ENZ-52309, Tetramethylrhodamine ethyl ester perchrolate, Enzo Life Sciences, Exeter, UK) for 30 minutes at 37°C. Positive control cells were incubated with 20 nM FCCP (ab120081, Abcam) for 5 minutes at 37°C prior to staining. After staining, cells were rinsed with prewarmed PBS and analyzed with flow cytometry on MACSQuant Analyzer 10 (Miltenyi Biotec, Bergisch Gladbach, Germany). Median fluorescence intensity (MFI) was measured using FlowLogic 8.6 software (Inivai Technologies, Mentone, Australia).

### Western blotting

Total protein or mitochondrial fractions from lymphoblasts were lysed, separated by SDS-PAGE and transferred to blotting membranes as described.^28^ Membranes were incubated with the following primary antibodies: anti-COX18 (Protein atlas, HPA049489, 1:1000), anti-MTCO2 (Abcam, ab913117, 1:1000), anti-αTubulin (Abcam, ab7291, 1:5000), anti-VDAC1 (Abcam, ab14734,1:1000), anti-SDHA (Genetex, GTX632636, 1:3000) anti-HCCS (Proteintech, 15118-1-AP, 1:2000), OXPHOS Human WB Antibody cocktail (Abcam, ab110411, 1:200), anti-ATP5C1 (ThermoFisher Scientific, 60284-1-IG, 1:1000). Anti-rabbit IgG (Promega, W401B, 1:10000), anti-mouse IgG1 and IgG2b (Southern Biotech, 1070-05, 1090-05, 1:10000) were used as secondary antibodies. The blots were developed with Pierce ECL Plus substrate (Thermo Fisher Scientific) and imaged using the Amersham Imager 600 (GE Healthcare, Wauwatosa, WI, USA). Images were analyzed using ImageJ^34^ to calculate the mean pixel gray values of each band.

### Drosophila stocks and maintenance

The following fly stocks were obtained from the Bloomington Drosophila Stock Center (Bloomington, IN, USA): UAS-mCD8.ChRFP (BL27391), dpr-Gal4 (BL25083), Mi{MIC}CG4942^MI03165^ (dCOX18^+/-^, BL36211). The RNAi line for Sply (RNAi Sply) was obtained from Vienna Drosophila Resource Center (Vienna, Austria) (Sply^HMS02526^, v42834). The UAS-YARS1 fly line expressing human YARS1 with a CMT-causing variant (YARS1-E196K) was generated and described previously.^35^ The nsyb-Gal4 driver line was kindly provided by M. Leyssen and B. Dickson.^36^ All crosses were performed at 25°C, 12h light/dark cycle, on Nutri-Fly™ flood (Flystuff; San Diego, USA).

### Fly RNA extraction, cDNA synthesis and RT-qPCR

Total RNA was isolated from the heads of adult flies following standard Trizol (Qiagen, Hilden, Germany) and chloroform extraction protocol. RNA was treated with TURBO DNA-free kit (Invitrogen, Thermo Fisher Scientific, Waltham, MA, USA) followed by cDNA synthesis using iScript Advanced cDNA synthesis kit for RT-qPCR (Bio-Rad). To confirm the downregulation of *COX1*8’s ortholog, dCOX18 (CG4942), in the heterozygous null fly line, gene expression levels were determined by quantitative RT-PCR using SYBR green PCR master mix (Applied Biosystems, Waltham, MA, USA) and compared to a control *yw* fly line. The following targets were used as housekeeping genes: Gapdh1, RpL32, RpS13, Act42A, Act79B. The housekeeping gene with the most stable expression across samples was used to normalize the data. The results were analyzed using the qBase+ software (Biogazelle, Gent, Belgium). The primers used for RT-qPCR are described in the Supplementary Table 2.

### Wing degeneration assay

A *Drosophila* wing degeneration assay was performed as described previously.^37^ Flies carrying the UAS-mCD8ChRFP construct in their genome were crossed with flies with the dpr-Gal4 driver to express the mChrerry protein in the chemosensory neurons of the wing margin bristles. The construct encodes a membrane-bound RFP-labeled mCherry protein that is incorporated into the neuronal membrane. When the flies reached the desired age (1 day or 30 days post eclosion), one wing per fly was clipped as close as possible to the thorax. Each wing was washed with PBS supplemented with 0,2% Triton X-100 and then mounted in Dako mounting medium (Agilent technologies, Santa Clara, CA USA). Axonal degeneration was determined by visual inspection of the fragmentation of the neuronal membrane and the proportion of wings depicting the fragmented phenotype per fly line was calculated.

### Negative Geotaxis Assay

The effect of dCOX18 downregulation on fly locomotion was studied using a negative geotaxis assay as described.^38^ From each fly line, 10 days post eclosion female flies with clipped wings were placed in a closed vial of 49 mm diameter. Fly movements were recorded using an infrared camera. After acclimation for 1 hour, the flies were tapped onto the bottom of the vial using a semi-automated FlyCrawler device (Peira Scientific Instruments, Beerse, Belgium). For each genotype, 10 groups of 10 flies were tested at least 15 times and the average time taken by the 10 fastest flies to reach a mark at a height of 82 mm was calculated.

### Statistical analyses

GraphPad Prism 9.2.0 was used for statistical analyses. The test for each experimental measurement is reported within the figure legends. Briefly, continuous variables were compared with a two-tailed Student’s t-test or a one-way ANOVA followed by Tukey’s multiple-comparison test. Categorical variables were analyzed by pairwise comparison with a one-sided Chi-square test. In all figures p-values are reported as * *P*<L0.05, ** *P*<0.01, *** *P*<0.001, **** *P*<0.0001, and *ns* for non-significant.

## Data availability

Data generated during this study can be shared by the corresponding author on reasonable request from any qualified investigator.

## Results

### Exome sequencing identifies three independent families with biallelic deleterious variants in *COX18*

ES was performed in the affected siblings from Family 1. First, we screened for potential pathogenic variants in known CMT genes, but no causal variants were found. We then conducted a HOMWES analysis which revealed 17 regions of homozygosity shared between both affected individuals, totaling 33.4Mb in size (the biggest of 8Mb). Variant filtering within those regions and prioritization based on impact and population frequency led to the identification of a homozygous splice variant NM_001297732.2:c.435-6A>G in the second intron of *COX18*. The variant was confirmed by Sanger sequencing and analysis of available relatives showed that it co-segregated with the disease (Fig. 1A). The variant was extremely rare in the control population database gnomAD v4.1.0^39^, with an allele frequency of 0.00001 and no homozygotes. The variant was predicted by multiple in silico tools (SpliceSiteFinder-like^40^, MaxEntScan^41^ and NNSPLICE^42^) to abolish the canonical acceptor site in exon 3 and to generate a new acceptor site 6 bp upstream in intron 2 (Fig. 2A). The resulting frameshift was therefore expected to create a premature stop codon in exon 3 and to cause nonsense-mediated decay of *COX18* canonical transcripts (Fig. 2A).

**Figure 1.**
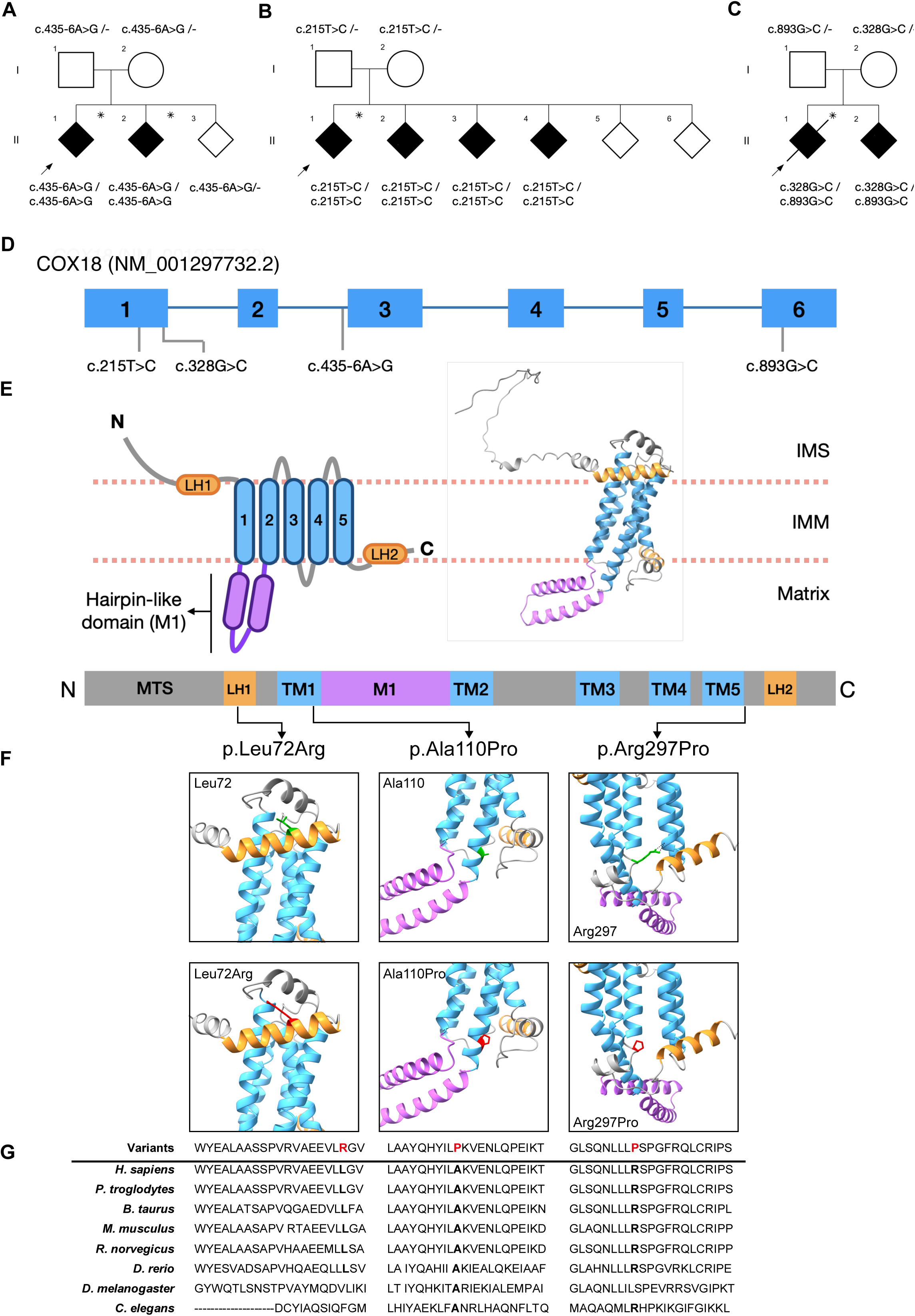
Biallelic variants in *COX18* in three families with autosomal recessive axonal CMT. (**A**) Pedigree and variant segregation of the Families 1-3. Squares indicate males and circles represent females. Black symbols indicate affected individuals. Arrows point at the proband of each family. Asterisks specify the patients whose exome was sequenced. The diagonal line across a symbol denotes a deceased individual. (**B**) Distribution of the variants on the COX18 gene (top) and protein (bottom). Diagram of COX18 domains (middle left) and predicted protein structure model of COX18 (middle right) (**C**) Amino acid conservation of the Leu72, Ala110, and Arg297 residue across multiple species. (**D**) Position of affected residues in green (top panels) and mutations in red (bottom panels) in the predicted COX18 protein structure. IMS = intermembrane space; IMM: inner mitochondrial membrane; MTS: mitochondrial targeting sequence; LH: amphipathic helix; TM: transmembrane helix; M1: hairpin-like domain.

**Figure 2.**
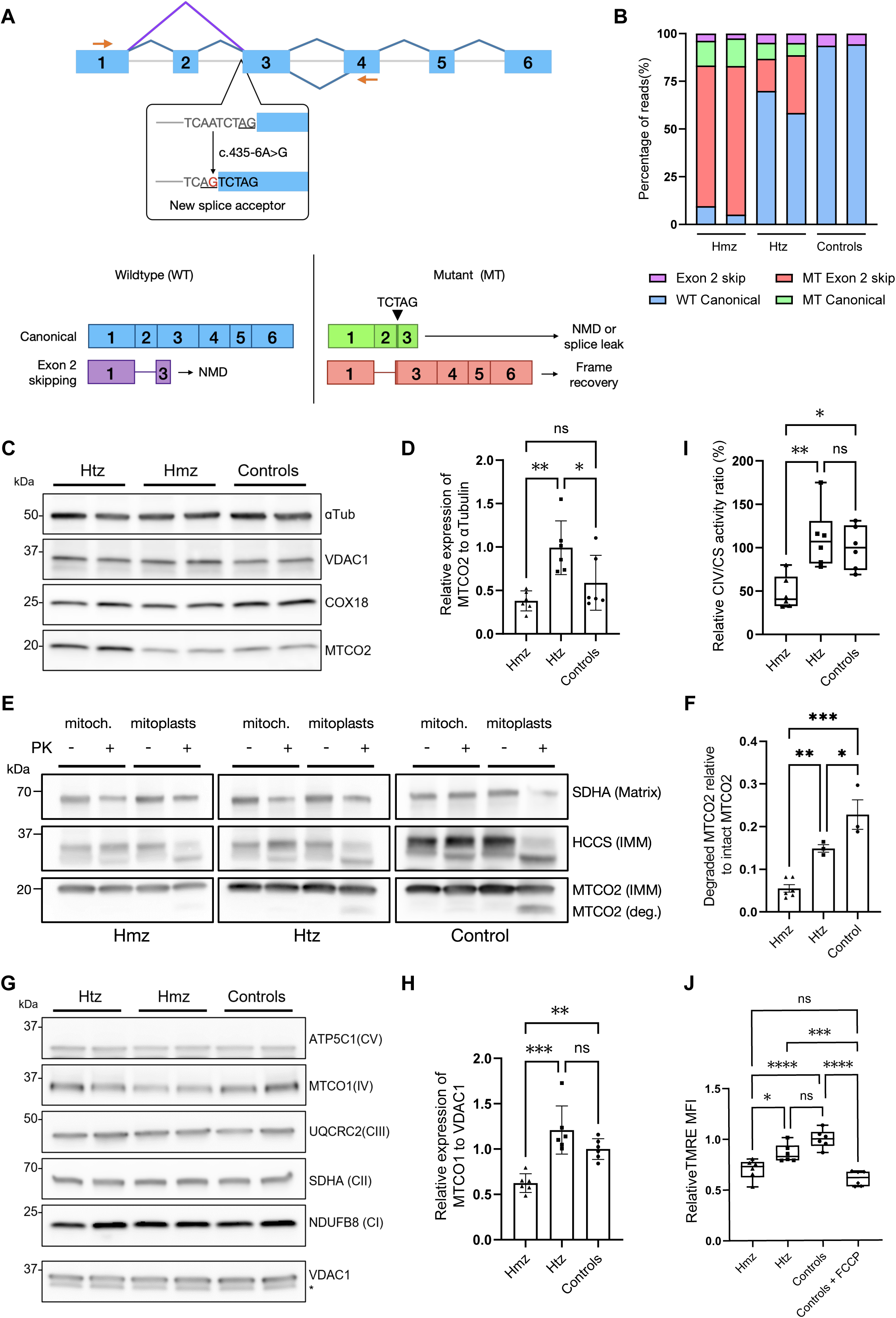
Functional characterization of COX18:c.435-6A>G variant in patients’ EBV-transformed lymphoblasts. (**A**) Exon-intron architecture of wildtype *COX18* transcripts. Splicing junctions are depicted as carets connecting the exons, junctions of the canonical transcripts (ENST00000507544.3, ENST00000295890.8) are colored in blue and the junction of the exon 2 skip transcript (ENST00000449739.6) is shown in purple. Orange arrows represent the forward and reverse primers used for *COX18* cDNA T-LRS. The effect of the variant on splicing is shown inside a dialog box underneath exon 3. Diagrams of the wildtype transcripts are shown in the bottom left panel and the abnormally spliced transcripts that result from the splice mutation in the bottom right. (**B**) Relative quantification of *COX18* transcripts sequenced by cDNA T-LRS in homozygous (hmz), heterozygous (htz), and control individuals shows that the mutant transcript skipping exon 2 is the predominant transcript in the homozygous patients (n = 1 for each genotype, with 2 biological replicates for each genotype). (**C** and **D**) Immunoblotting of full lysate protein from EBV-transformed lymphoblasts derived from the 3 genotypes shows no reduced levels of COX18. Western blot analysis revealed reduced levels of MTCO2 in the homozygotes compared to the carrier but not to controls. Data are shown as mean ± SD (n = 3 for each genotype, with 2 biological replicates for each genotype). (**E** and **F**) Proteinase K (PK) protection assay indicates that in mitoplasts from lymphoblasts of the homozygous proband supplemented with proteinase K MTCO2 is protected from degradation, while heterozygotes and controls show increased levels of degraded MTCO2. The levels of degraded MTCO2 (deg. MTCO2) relative to the levels of intact MTCO2 in the untreated mitochondrial fraction are shown as mean ± SD (n = 3 for each genotype, 2 biological replicates were tested for the homozygous genotype). SDHA is a mitochondrial matrix protein and HCCS is an inner mitochondrial (IMM) protein. (**G** and **H**) Western blot of mitochondrial fractions from lymphoblasts revealed decreased protein levels of complex IV (CIV) subunit MTCO1 in the homozygotes relative to controls and heterozygotes. The asterisk indicates an unspecific band. Immunoblotting quantification of MTCO1 is shown as mean ± SD (n = 3 for each genotype, with 2 biological replicates for each genotype). (**I**) Normalized CIV to citrate synthetase (CS) activity is decreased in homozygotes compared to the other genotypes. Values are shown as mean ± SD (n = 3). (**J**) Flow cytometry analysis of mitochondrial membrane potential with TMRE staining shows decreased median fluorescence intensity (MFI) in lymphoblasts from homozygous patients relative to controls. Control lymphoblasts treated with FCCP, which depolarizes the mitochondrial membrane, were used as positive control. Data are shown as mean ± SD (n = 3 for each genotype, with 2 biological replicates for each genotype). \*\*\*\**P*<0.0001, \*\*\**P*<0.001, \*\**P*<0.01, \**P*<0.05, ns: not significant.

Screening for additional individuals with *COX18* biallelic mutations in our in-house cohort of 272 recessive and 90 sporadic unsolved CMT patients did not return any additional affected individuals. However, three additional families were found through the GENESIS^26^ and RD-Connect^27^ online platforms. Family 2 consisted of four siblings who harbored a homozygous NM_001297732.2:c.215T>G (p.Leu72Arg) (Fig. 1B). The variant is extremely rare (allele frequency of 0.00002, with no homozygotes in gnomAD.^39^ Family 3 carried compound heterozygous variants NM_001297732.2:c.328G>C (p.Ala110Pro) and c.893G>C (p.Arg297Pro) which were confirmed to be in trans (Fig.1C). Both variants are exceedingly rare with allele frequencies of 0.000001 and 0.00002, respectively, and had no homozygotes in gnomAD.^39^ All these missense variants co-segregated with the disease and were predicted to be deleterious by Polyphen-2 and CADD (CADD PHRED > 20).

All the substitutions affected highly conserved amino acids across different vertebrate species (Fig. 1G). As no crystal structure is available for COX18, AlphaFold^43,44^ was used to determine the location and potential impact of the substituted residues. Leu72 lies in a short amphipathic helix (LH1) that faces the inner mitochondrial space, parallel to the inner mitochondrial membrane (Fig.1E and F). The substitution by an arginine introduces a positive charge that might impact the hydrophobic interactions of that helix (Fig. 1F, *left panels*). Ala110 is at the matrix end of the 1^st^ transmembrane helix (TM1) (Fig. 1E and F, *middle panels*) and the introduction of a buried proline in its place is predicted to be structurally damaging by Missense 3D-SB^45,46^ (Fig. 1F, *bottom middle panel*). Arg297 residue is located at a loop just after the 5^th^ transmembrane helix (TM5) (Fig. 1E) and shares hydrogen bonds with Ile241 in the 3^rd^ transmembrane domain (Fig. 1G, *top right panel*). The Arg297Pro substitution is predicted by Missense 3D-SB^45,46^ to damage the protein structure due to disallowed conformations. Additionally, we hypothesize that the change might abolish the hydrogen bond shared with Ile241, a residue from the 3^rd^ transmembrane helix (TM3) (Fig. 1G, *bottom right*), which might be key for the protein folding. This is supported by FoldX^47^ in silico tool which estimated that the Arg297Pro could result in a change of protein stability of 3.8kcal/mol.

### *COX18* deficiency causes a predominant sensory-motor neuropathy phenotype

Detailed clinical features of all nine affected individuals are described in Table 1. The cardinal feature among all patients was progressive axonal polyneuropathy with a variable onset, ranging from infancy to adulthood. All patients initially presented with distal lower limb symptoms and, as the disease progressed, most of them slowly developed distal upper limb involvement. Most of the patients presented muscle weakness at disease onset, except for two siblings from family 2, whose phenotypes are predominantly characterized by sensory loss in distal lower limbs. Muscle atrophy was prominent in the feet, the tibialis anterior and the calf muscles. Two of the patients were dependent on mobility aids, such as walking stick or wheelchair. All patients presented sensory loss which followed a stocking and glove pattern. The sense of touch, proprioception and vibration were impaired, mostly in the lower limbs. One of the individuals (3.II.1) in addition developed, from their mid-thirties, pain in feet and thighs accompanied by a band-like pressure sensation around the legs. Their affected sibling (3.II.2) also complained about mild pain until the proximal segment of the calf. All patients suffered from foot deformities, usually in the form of pes cavus, but some of them also presented pes equinovarus and hammer toes. Most of the patients exhibited decreased or absent deep tendon reflexes in distal lower limbs except for the patients from family 3. More specifically, individual 3.II.1 had generalised brisk reflexes except for absent ankle reflexes and as the disease progressed their gait became increasingly spastic and ataxic. Their sibling (3.II.2) had a similar presentation with general hyperreflexia and absent ankle reflexes on examination.

**Table 1.**
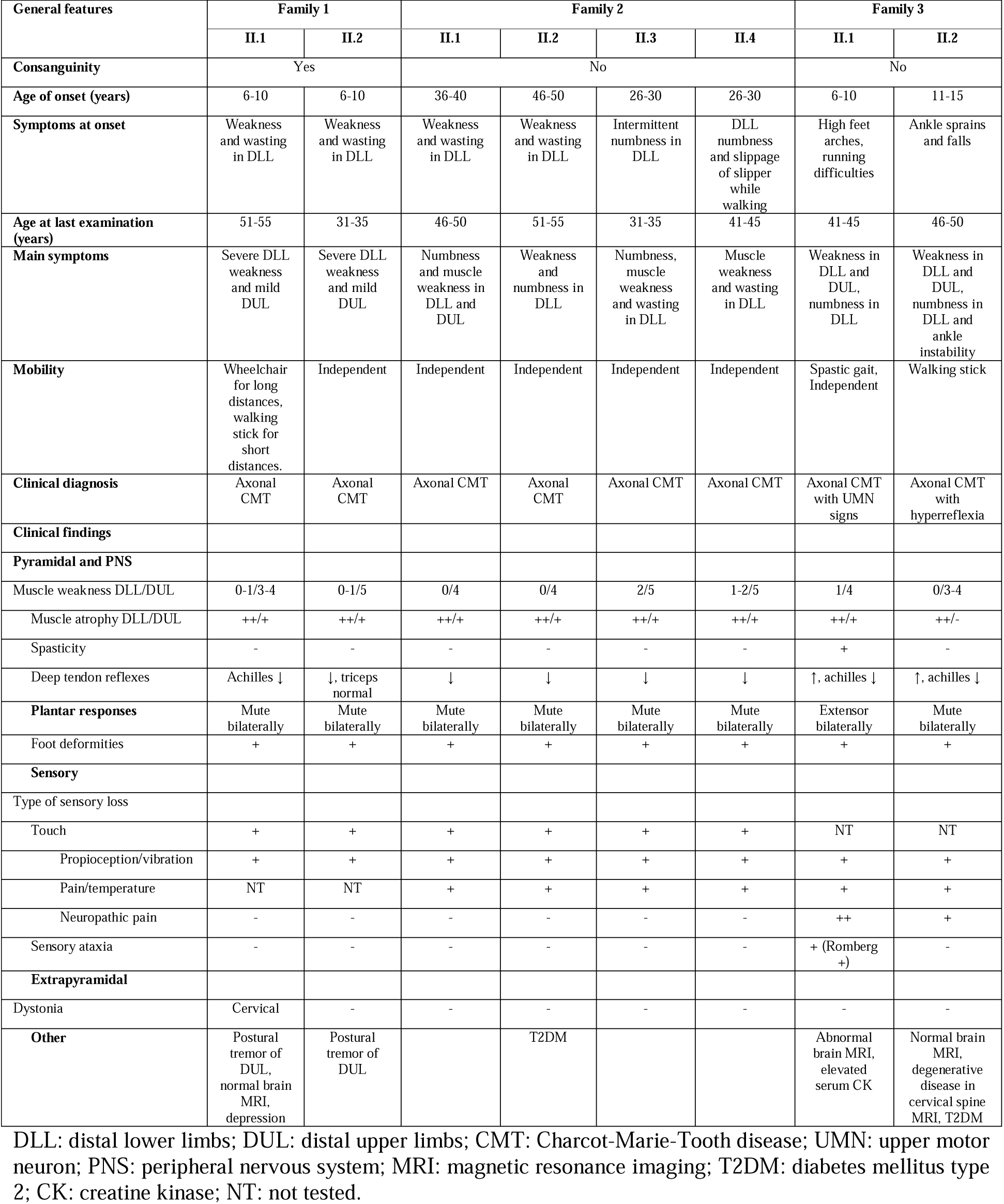
Clinical features of the index patients.

Nerve conduction studies (Table 2) revealed decreased compound muscle action potential (CMAP) in peroneal nerve recordings and, in some patients, in the median nerve recordings as well. Sensory nerve action potentials (SNAP) were mostly undetectable in both ulnar and sural nerves. Motor nerve conduction velocity (mNCV) was within normal ranges in most patients. Some of the individuals exhibited slightly decreased mNCV which in the context of low amplitudes was attributed to axonal loss. In summary, electrophysiologic studies indicated sensory-motor axonal neuropathy in all the patients.

**Table 2.** Electrophysiological studies of the index patients.

| Nerve | NCS | Family 1 |  | Family 2 |  |  |  | Family 3 |  |
| --- | --- | --- | --- | --- | --- | --- | --- | --- | --- |
|  |  | II.1 | II.2 | II.1 | II.2 | II.3 | II.4 | II.1 | II.2 |
| Median | CMAP | 5.8 mV | 3.3 mV | 83 μV | 248 μV | 6.5mV | 350 μV | 7.6 mV | 6.4 mV |
|  | mNCV | 47.7 m/s | 55.9 m/s | 50 m/s | 36m/s | 58.2m/s | 30.0 m/s | 55 m/s | 48 m/s |
| Peroneal | CMAP | ND | ND | ND | 452 μV | 4.53 mV | ND | 1.2 mV | 0.8 mV |
|  | mNCV |  |  |  | 24.0 m/s | 40.6 m/s |  | 44 m/s | 28 m/s |
| Ulnar | SNAP | 23,2 μV | NP | 20.8 μV | ND | 30.5 μV | ND | ND | ND |
|  | sNCV | 40,0m/s |  | 28.6 m/s |  | 26.8 m/s |  |  |  |
| Sural | SNAP | ND | ND | ND | ND | ND | ND | ND | ND |
|  | sNCV |  |  |  |  |  |  |  |  |
| Electrophysiological diagnosis |  | Axonal CMT | Axonal CMT | Axonal CMT | Axonal CMT | Axonal CMT | Axonal CMT with secondary demyelinating changes | Axonal CMT | Axonal CMT |

NCS: nerve conduction studies; CMAP: compound muscle action potential; mNCV: motor nerve conduction velocity; SNAP: sensory nerve action potential; sNCV: sensory nerve conduction velocity; ND: not detected; NP: not performed; CMT: Charcot-Marie-Tooth disease; NA: not applicable.

During the disease course, some patients developed additional symptoms beyond the peripheral nervous system. Affected individuals from family 1 had upper limb tremor and individual 1.II.1 developed cervical dystonia around 41-45 years of age, which was treated with botulinum toxin. Patients from family 3 showed brisk deep tendon reflexes and individual 3.II.1 had a particularly complex phenotype characterized by sensory ataxia, spastic gait, and bilateral positive Babinski reflexes. Follow-up studies in this patient revealed elevated serum creatine kinase 165 IU/L (normal range 26-140 IU/L) and brain and spinal MRI revealed mild cerebellar atrophy and spinal degenerative changes. Their affected sibling (3.II.2) showed a normal brain MRI and degenerative diseases with C5-6 exit foraminal compromise. Noteworthy, none of the patients presented other mitochondrial-related features such as cardiomyopathy, hepatopathy, and tubulopathy.

### Splice variant c.435-6A>G leads to alternative splicing resulting in expression of an aberrant COX18 isoform

To identify the impact of the c.435-6A>G variant on splicing, cDNA T-LRS was carried out to reliably identify and quantify the different *COX18* transcripts (Fig. 2A and 2B). To do this, *COX18* cDNA was amplified from EBV-transformed lymphoblasts from the patients, their parents, and unrelated controls. Primers were designed in exon 1 and 4 (Fig. 2A, Supplementary Table 1) to be able to distinguish all previously known and potentially novel *COX18* transcripts and their associated splicing events. Sequencing confirmed that the splice variant generated a new acceptor site in intron 2 that added five coding base pairs to exon 3 (Fig. 2A). The frameshift caused by this insertion generated a premature stop codon in the third exon of the canonical transcripts (Ensembl IDs ENST00000507544.3 and ENST00000295890.8). Accordingly, the mutant canonical transcripts represented a minor portion of the reads in patients and carriers, which suggests that they are most likely degraded by non-sense mediated mRNA decay (NMD) (Fig. 2B). Interestingly, an alternative transcript skipping exon 2 represented as much as 66% of all the transcripts in the patients and 14-27% in the heterozygotes (Fig. 2B). This transcript corresponds to an alternative transcript lacking exon 2 (ENST00000449739.6) that has a premature stop codon in exon 3 and is normally degraded by NMD. However, instead of being degraded, this transcript recovers its reading frame as a consequence of the aberrant splicing. Canonical wildtype *COX18* transcripts (ENST00000507544.3 and ENST00000295890.8) totaled 86% of the transcripts in controls and 53-61% in heterozygote carriers (Fig. 2B). In contrast, these canonically spliced transcripts only constituted 4-8% in patients, suggesting that splicing leakage occurs at negligible levels. Another NMD transcript (ENST00000510031.1) with 4bp added to exon 2 was observed in all samples in a very small proportion, regardless of the genotype (data not shown).

### Mutant COX18 protein affects the assembly and stability of CIV subunits

COX18 is an assembly factor that translocates the C-terminus of MTCO2, a core subunit of CIV, across the inner mitochondrial membrane (IMM) into the inner mitochondrial space (IMS). Models in yeast and human cells have demonstrated that *COX18* knock-out (KO) impairs MTCO2 insertion across IMM and renders it unstable, ultimately leading to MTCO2 degradation.^48,49^ To evaluate the impact of the splice mutation on the quantity and function of COX18 and its partner MTCO2, we performed immunoblotting assays with patients’ lymphoblasts. Western blotting showed that COX18 is stably expressed in the patients in comparable amounts to controls (Fig.2C, Supplementary Fig. 2). However, MTCO2 levels were decreased in patients compared to their heterozygous parents, but not to controls (Fig.2C and D), suggesting that mutant COX18 might affect MTCO2 protein levels. We further examined COX18’s ability to translocate MTCO2 across the IMM using a proteinase K (PK) protection assay. The experiment showed that MTCO2 was PK-sensitive and degraded in mitoplasts from the controls and the carriers, while in the patients’ mitoplasts it was protected from degradation (Fig.2E and F). These results point out that mutant COX18 has an impaired ability to insert MTCO2 C-terminus across the IMM, affecting its stability and expression level. To assess the impact of mutant COX18 on the stability of CIV or other complexes, a representative subunit from each complex was immunoblotted (Fig. 2G). The experiment revealed decreased levels of CI subunit MTCO1, while the subunits from other complexes did not show altered protein levels (Fig. 2H). This suggests that the variant may be associated with an isolated defect in CIV, but further follow-up studies are needed to confirm this.

### Splice mutation c.435-6A>G is associated with decreased CIV activity and mitochondrial membrane potential in patient-derived lymphoblasts

Considering that COX18 loss of function affects the stability of a core subunit of CIV, we evaluated whether this defect might impact its overall enzymatic activity. CIV is the final enzyme from the electron transport chain. It catalyzes the oxidation of reduced cytochrome c which generates water. This reaction is coupled with the transport of four protons across IMM, contributing to the proton gradient that drives ATP synthesis. To assess the impact of the variant on CIV activity, we performed an enzymatic assay that measures the rate of change in absorbance at 550nm caused by the oxidation of cytochrome c. The assay revealed that patients had a lower CIV to citrate synthetase (CS) enzymatic activity ratio compared to controls (Fig. 2I). In turn, decreased CIV activity could lead to reduced proton translocation through the IMM. Therefore, mitochondrial membrane potential was measured using flow cytometry on patients’ lymphoblasts stained with TMRE, a dye that accumulates in cells with hyperpolarized IMM. TMRE signal was significantly lower in the probands cells compared to carriers and controls, which indicates a decrease in mitochondrial membrane potential (Fig. 2J).

### *Drosophila melanogaster* COX18 knockdown model displays signs of neurodegeneration

COX18 is a key assembly factor of the mitochondria, and as such, has proven to be functionally conserved from fungi to mammals. ^50,51^ *COX18* ortholog in *Drosophila melanogaster* (CG4942, dCOX18) shares 61% similarity and 40% identity to the human protein (Supplementary Fig. 1).^52^ No fly phenotype has been reported to be associated with dCOX18 downregulation. To emulate the loss of function of *COX18* observed in the patients, we used a fly line with one copy of dCOX18 disrupted by the insertion of the transposon Minos-mediated integration cassette (MiMIC) within its coding sequence. Notably, this leads to around 90% reduction of dCOX18 mRNA levels in comparison to naïve flies (yw, Fig. 3B), while pan-neuronal downregulation of dCOX18 expression only led to approximately 75% reduction (Nsyb dCOX18-RNAi, Fig. 3B). Furthermore, when crossing dCOX^+/-^ flies to obtain homozygous dCOX18-deficient flies, we observe no offspring, suggesting that the complete loss of dCOX18 in the homozygous null flies is not viable. Therefore, we chose the dCOX18^+/-^ fly line as it was the most biologically relevant model for the recessive disease we could to study.

**Figure 3.**
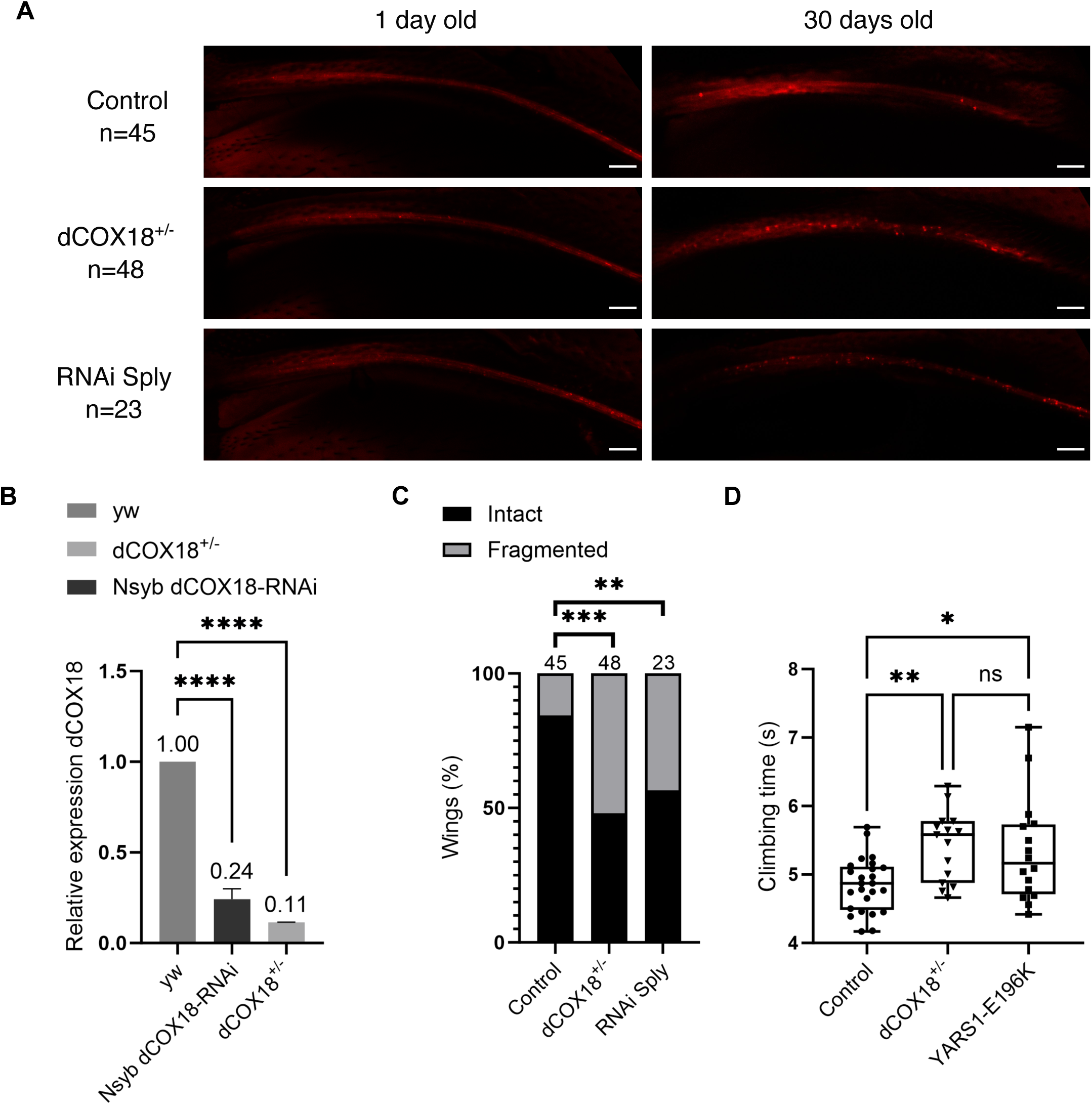
Downregulation of *COX18*’s ortholog in Drosophila, dCOX18, causes age-dependent axonal degeneration of sensory neurons and locomotor impairment. (**A**) The nerve tract along the L1 wing vein was visualized by mCherry expression using dpr-Gal4 driver. Representative images are shown from 1-day and 30-day-old flies from each genotype. RNAi Sply flies were used as positive control. Flies expressing the driver alone were used as negative control. (**B**) Quantification of dCOX18 RNA expression using RT-qPCR for control (yw), Nsyb-driven RNAi dCOX18 pan-neuronal knockdown (Nsyb dCOX18-RNAi), and heterozygote null (dCOX18^+/-^) fly lines. Data are shown as mean ± SD (n=2). (**C**) Quantification of the percentage of wings with axonal fragmentation at day 30. (n = 23-48 per genotype). (**D**) Climbing performance of control (yw), dCOX18^+/-^, and YARS1-E196K fly lines was assessed by measuring the climbing time of 10-day-old flies. Data are shown as mean ± SD (n = 15-25, 10 groups of 10 flies tested for each phenotype). \*\*\*\**P*<0.0001, \*\*\**P*<0.001, \*\**P*<0.01, \**P*<0.05, ns: not significant.

We assessed then whether the heterozygote dCOX18^+/-^ displayed behavioral and histopathological signs of neurodegeneration. First, we performed a wing degeneration assay to assess if dCOX18 partial loss is associated with axonal degeneration. We evaluated the integrity of the long axons of the chemosensory neurons innervating the wing margin bristles, by expressing a fluorescent red mCherry protein (UAS-mCD8chRFP) in the neuronal membrane (dpr-Gal4 driver) (Fig. 3A). No difference was observed between the fly lines carrying different genotypes on the first day after eclosion. Nonetheless, 30-day-old dCOX18^+/-^ flies exhibited prominent axonal fragmentation compared to the control fly line (Fig. 3A and C). This degenerative phenotype was comparable to the effect caused by downregulation of Sply, the *Drosophila* ortholog of *SGPL1* that causes axonal CMT in humans.^37^ In addition, a negative geotaxis climbing assay was conducted to evaluate the locomotor performance of the dCOX18^+/-^ flies (Fig. 3D). In this test, the dCOX18^+/-^ flies presented a slower climbing speed in comparison to the controls (Fig. 3D). The dCOX18^+/-^ flies climbing performance was similar to the locomotion deficit presented by another well-established *Drosophila* model, expressing a CMT-causing mutation in the *YARS1* gene.^53^

## Discussion

This study provides genetic and functional evidence to support *COX18*, a nuclear-encoded mitochondrial assembly factor, as a novel CMT gene candidate. Biallelic missense and splice variants in *COX18* were identified in three families with autosomal recessive axonal CMT.

*In-silico* predictions, *in vitro* and *in vivo* studies, suggest that *COX18* loss of function is the underlying disease mechanism. Congruently, downregulation of the *COX18* homolog in *Drosophila* replicates key features of neurodegeneration, such as locomotor impairment and axonal degeneration of sensory neurons.

After screening in-house and external CMT cohorts, we have found in total eight patients from three families with biallelic variants in *COX18*. All patients showed sensory-motor axonal polyneuropathy that primarily affects the distal lower limbs. They presented with muscle weakness and atrophy accompanied by foot deformities. The motor impairment significantly affected the ambulation of the patients from families 1 and 2, who were dependent on mobility aids. All individuals experienced sensory loss which was more predominant amongst the affected members from family 3. Electrophysiological studies revealed axonal degeneration of motor and sensory nerves from upper and lower limbs. This is compatible with the literature as the most common type of neuropathy observed in mitochondrial disorders is axonal.^54–56^

Peripheral neuropathy occurs in approximately one-third of the patients with mitochondrial diseases.^57–61^ As mitochondrial function is essential for many tissues and systems, mitochondrial peripheral neuropathy usually occurs together with other neurological and extra-neurological manifestations, such as encephalopathy, myopathy, cardiac disease, renal dysfunction ^56,57,59^. In some cases, neuropathy can be the only manifestation at onset, but subsequently other tissues might become affected as the disease progresses.^57,62,63^ Likewise, some of the patients reported here developed CNS symptoms during the course of the disease. For instance, patient 1.II.1 developed cervical dystonia in their 40s, three decades after the disease onset. Family 2 seems to be the exception, as all four affected siblings do not show any CNS manifestation. Nevertheless, their peripheral neuropathy has a late onset (^~^40s), and it is still plausible that additional symptoms might develop at later stages of the disease.

COX18-related neuropathy showed notable inter-and intra-familial phenotypic variability. Disease onset varied considerably between families, from late childhood to middle adulthood. This clinical heterogeneity might be due to the distinct impact of each variant on COX18 function or the differences in genetic background and epigenetics between the patients. Mitochondria are highly dynamic organelles with the ability to alter their mass, shape, and number to compensate metabolic insults.^64^ Moreover, due to the dual genetic origin of mitochondrial proteins, the interaction between nuclear (nDNA) and mitochondrial DNA (mtDNA) might modulate the penetrance and severity of mitochondrial disease.^65–67^ For example, mtDNA variants have been shown to significantly influence the phenotype of mice with mutations in mitochondrial nDNA genes causing cardiomyopathy.^68^ Finally, environmental modifiers might also play a role in determining the severity and progression of mitochondrial disorders, as observed in mitochondrial optic neuropathies.^69,70^

In this study, we have identified four different *COX18* variants, three missense and one splice variant. All were segregated with the disease in each family and were either novel or extremely rare in public databases. All three missense variants perturbed conserved residues in different domains of the protein. In silico predictions suggest that they are likely deleterious. The p.Ala110Pro and p.Arg297Pro variants are predicted to disrupt the protein structure. While p.Leu72Arg might not affect COX18 structure, it introduces a positive charge in the LH1 amphipathic helix that might affect its function. This domain has been proven to be essential for the insertase function of proteins from the same family (Oxa1/YidC/Alb3) and is thought to destabilize the lipid bilayer and facilitate the release of the inserted protein into the membrane.^71,72^ Functional validation of the missense variants is necessary to understand the precise effect of the predicted structural perturbations on COX18 activity.

We studied in detail the functional effect of the splice variant c.435-6A>G. The variant reduces the expression of the canonical transcript to negligible levels, and at the same time, generates an alternatively spliced product missing exon 2. This aberrant transcript was demonstrated to lead to the expression of a stable but partially functional mutant protein. Consistent with our findings, deletions in the same region in the *COX18* ortholog, *yidC*, in *Bacillus subtilis* and *Escherichia coli* do not affect the protein stability but significantly impair its translocase function.^72,73^ According to COX18 *in silico* structural models^43,44^, the loss of exon 2 would disrupt a helical hairpin (M1) (Fig.1C) that is well conserved in all translocases from the Oxa1/YidC/Alb3 family.^74^ It is hypothesized that this dynamic and flexible hairpin is in charge of substrate recruitment^74,75^. Likewise, our results highlight the importance of this domain as the aberrant COX18 isoform identified shows a reduced ability to translocate MTCO2 C-terminus, probably owing to difficulties in its recruitment. MTCO2 is a core subunit of CIV which accepts the electrons from cytochrome c through the copper center in its C-terminal and transfers them across the complex to produce water.

Patients with the homozygous splice variant showed decreased levels of COX18’s substrate MTCO2 compared to their heterozygote parents. Yet, the affected individuals expressed similar MTCO2 protein levels than controls. This could be a result of an increase in MTCO2 protein synthesis as a compensatory mechanism. Furthermore, the PK protection assay in mitoplasts from the patients revealed that MTCO2 was not properly inserted in the IMM. Therefore, it is likely that the decreased MTCO2 levels that we see in the patients are due to defects in the insertion and folding of MTCO2 which render it unstable and prone to degradation. These findings are supported by previous studies in *COX18* KO HEK293T cells which demonstrated that COX18 translocase activity is necessary for the post-translational stability of MTCO2.^49^ The MTCO2 deficiency observed in the patients was also associated with decreased levels of another core CIV subunit, MTCO1, suggesting a deleterious effect on the stability of other CIV subunits. In turn, these defects correlated with reduced CIV enzymatic activity and impaired mitochondrial membrane potential. Taken together, we demonstrate that the splice mutation affects the chaperone and translocation function of COX18, which ultimately affects the role of CIV as part of the mitochondrial electron transport chain.

*COX18* is expressed in multiple tissues, with the highest expression in EBV-transformed lymphoblasts, adrenal glands, and peripheral nerves.^76^ To assess the susceptibility of neurons to *COX18* downregulation in a whole organism, we studied a dCOX18 deficient fly model. The dCOX18^+/-^ fly displayed signs of neurodegeneration, including axonal degeneration of sensory neurons in the wing and age-dependent locomotor impairment. These results were comparable to the phenotypes observed in previously published CMT fly models, studying mitochondrial or non-mitochondrial CMT-associated genes, which exhibit axonal degeneration and climbing defects as the dCOX18^+/-^ fly.^37,38,77,78^ A similar locomotor impairment has been described in a fly knockdown model of *COA7*, another assembly factor of CIV that has been reported to cause CMT.^78^ Additional functional studies on *in vivo* and *in vitro* neuronal models are required to understand the susceptibility of the motor and sensory neurons to the loss of COX18 and to CIV deficiency in general.

A homozygous null *COX18* fly was not possible to obtain, which might suggest that *COX18* complete KO is not viable. While full KO of this gene may be lethal, flies expressing ^~^10% of COX18 ortholog, were viable and fertile, and demonstrated neurodegenerative phenotypes. Similarly, *COX18*^-/-^ mice exhibit embryonic growth retardation eventually leading to prenatal or preweaning lethality.^79^ It is worth noting that some of these mice show abnormalities in the neural tube closure. Altogether, a complete COX18 loss of function seems to be incompatible with life in different species. Therefore, we hypothesize that the variants reported in this study are probably hypomorphic and only partially reduce COX18 function or protein levels.

Despite an earlier study that screened a cohort of patients with CIV deficiency for *COX18* pathogenic variants,^80^ the gene has not been linked to any human disease until recently^81^. The first report of COX18-related pathology described a patient with neonatal encephalo-cardiomiopathy and CIV deficiency who carried a homozygous NM_001297732.2:c.667G>C p.(Asp223His) variant.^81^ Remarkably, this patient presented sensory-predominant axonal peripheral neuropathy as well. A recent publication identified through ES and homozygosity mapping *COX18* biallelic variants as a candidate cause for non-syndromic hearing loss, yet functional evaluation was not conducted.^82^ Thus, exhaustive phenotyping of additional patients is needed to ascertain the clinical spectrum of COX18-related conditions and further support its role in CMT pathogenesis.

Our findings on COX18-related CMT neuropathy illustrate that in the rare cases where peripheral neuropathy is the main or only clinical feature of an underlying mitochondrial disorder, it is likely to overlook the mitochondrial origin.^57,63^ The underlying mitochondrial etiology can be suspected through histochemical, biochemical, and neuroimaging studies. However, no single biomarker is sensitive enough to completely confirm the diagnosis and the lack of abnormal biomarkers does not exclude a mitochondrial dysfunction.^69,83^ Similarly, mitochondrial biomarkers, including serum lactate and MRI, did not show consistent findings suggestive of a mitochondrial pathology in our patients. The application of unbiased genetic testing was crucial for establishing the correct etiology. Likewise, using genetic approaches, several studies in the past decade have found different proteins of the mitochondrial respiratory chain to be implicated in the pathogenesis of CMT.^78,84–89^ Thus, our findings, together with these reports, stress the importance of screening mitochondrial genes as part of the diagnostic work-up of patients with CMT with or without a multisystemic clinical presentation.

Among them, biallelic mutations in genes encoding subunits or assembly factors of CIV particularly (*SURF1*, *COA7*, *COX6A1,* and *COX20*) have been found to cause CMT.^78,84–89^ Strikingly, some of these genes encode proteins that are also involved in the translocation and maturation of MTCO2. For example, COX20 plays a role as COX18’s counterpart by translocating the N-terminus of MTCO2.^87,90,91^ SCO2 is a metallochaperone that adds copper to MTCO2’s C-terminus once COX18 translocates it into the IMS.^84^ What is more, a missense mutation in the mitochondrial gene that encodes MTCO2, COX18’s substrate, has been reported to cause late-onset cerebellar ataxia, axonal peripheral neuropathy, and tremor.^92^ Interestingly, some of these genes have also been reported to cause severe multisystemic diseases, such as Leigh syndrome (*SURF1*) and cardioencephalomyopathy (*SCO2*).^93,94^ These reports, together with the findings of this study, underscore the role of CIV dysfunction in the pathogenesis of CMT. Further research is needed to understand what makes peripheral neurons particularly susceptible to CIV deficiency and to explain its broad clinical spectrum.

In conclusion, we have provided genetic and functional evidence to support COX18 as a new candidate gene for autosomal recessive axonal CMT. These findings underscore the importance of peripheral neuropathy in the spectrum of mitochondrial disorders, warranting the screening of mitochondrial genes in the diagnostic follow-up of CMT patients with or without CNS features. Our results also recommend the application of next-generation sequencing techniques in the diagnosis of non-syndromic neuropathies of mitochondrial etiology. Moreover, our study provides further evidence to support the critical role of the hairpin domain of COX18 for its translocase activity. Finally, we draw special attention to the impact of mitochondrial CIV deficiency in the pathogenesis of CMT.

## Acknowledgments

The authors acknowledge the VIB-UAntwerp Center for Molecular Neurology Neuromics Support Facility for their technical support. The authors also thank the patients and their families for participating in this study.

