## Supplementary Table 1 for "Biallelic variants in *COX18* cause a mitochondrial disorder primarily manifesting as peripheral neuropathy"

**Supplementary Table 1. Sequences of primers used in the study.**

| **Target** | **Forward** | **Reverse** |
| --- | --- | --- |
| **Targeted resequencing of *COX18* (NM_001297732.2)** | | |
| COX18 Exon 1 | GAAGGTCAATCGCGGCTG | TGATTAAACACTAAGCACTGCAG |
| COX18 Exon 2 | AGTTACTTGCACAGGATCACATG | CACACCATTGCACTCCAGAC |
| COX18 Exon 3 | GCAACTCCTTAACATGCCTG | GTTCACAACACAACCAATCTGT |
| COX18 Exon 4 | ACAGATTGGTTGTGTTGTGAAC | GAGAGGTTAAGGGACAGGCC |
| COX18 Exon 5 | CACTTGAGCCCAGGAGTTTG | CGGAGAATGGCGTGAACC |
| COX18 Exon 6 | TCCCAGGAATCTAGTAGCTGC | ACCTCGACCTCCCAAAGTG |
| **Targeted cDNA long-read sequencing of *COX18* (NM_001297732.2)** | | |
| Exon 1 to 4 | GCAGCATTCTGCTCTCCAC | ATTGATGACGCCAACAGAGA |
|  | Primers were supplemented with prefixes: | |
|  | TTTCTGTTGGTGCTGATATTGC | ACTTGCCTGTCGCTCTATCTTC |
| **Fly RT-qPCR** | | |
| Act79B | CAAGGATCTGTATGCCAACAATG | GGTCAGCGATACCTGGATACATG |
| Gapdh | CAGCCCCGACATGAAGGT | CGATCTCGAAGTTGTCATTGATG |
| RpS13 | GGGTCTGAAGCCCGACATT | GGCGACGGCCTTCTTGAT |
| RpL32 | GCTAAGCTGTCGCACAAATGG | CGGCGACGCACTCTGTT |
| Rp49 | TACAGGCCCAAGATCGTGAA | TCTCCTTGCGCTTCTTGGA |
| Act42A | CAGGCGGTGCTTTCTCTCTA | AGCTGTAACCGCGCTCAGTA |
| CG4942(dCOX18) | CCATGGCCAAGCACAAGTTC | GCTTCTTGATCGAACGCCGA |
