## Supplementary Figure 1 for "Biallelic variants in *COX18* cause a mitochondrial disorder primarily manifesting as peripheral neuropathy"

**Supplementary Figure 1.** Alignment Drosophila and human Cox18 protein sequences by Clustal Omega. The “*”symbol shows identical residues, the “:” indicates highly similar residues, the “,”shows residues with weak similarities, and no symbol indicates not conserved amino acids.

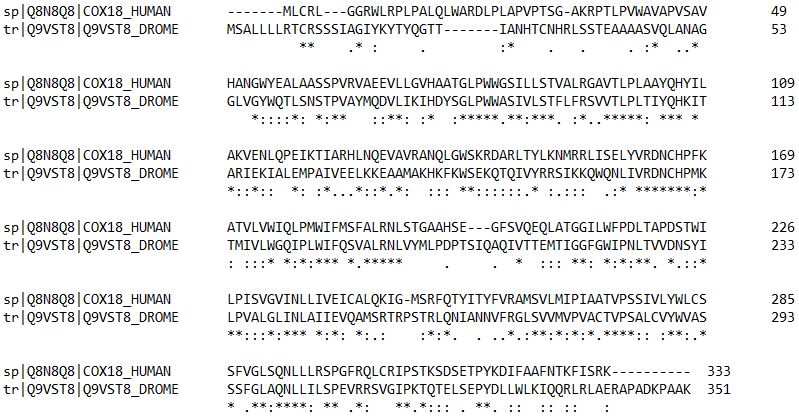
