## Supplementary Figure 2 for "Biallelic variants in *COX18* cause a mitochondrial disorder primarily manifesting as peripheral neuropathy"

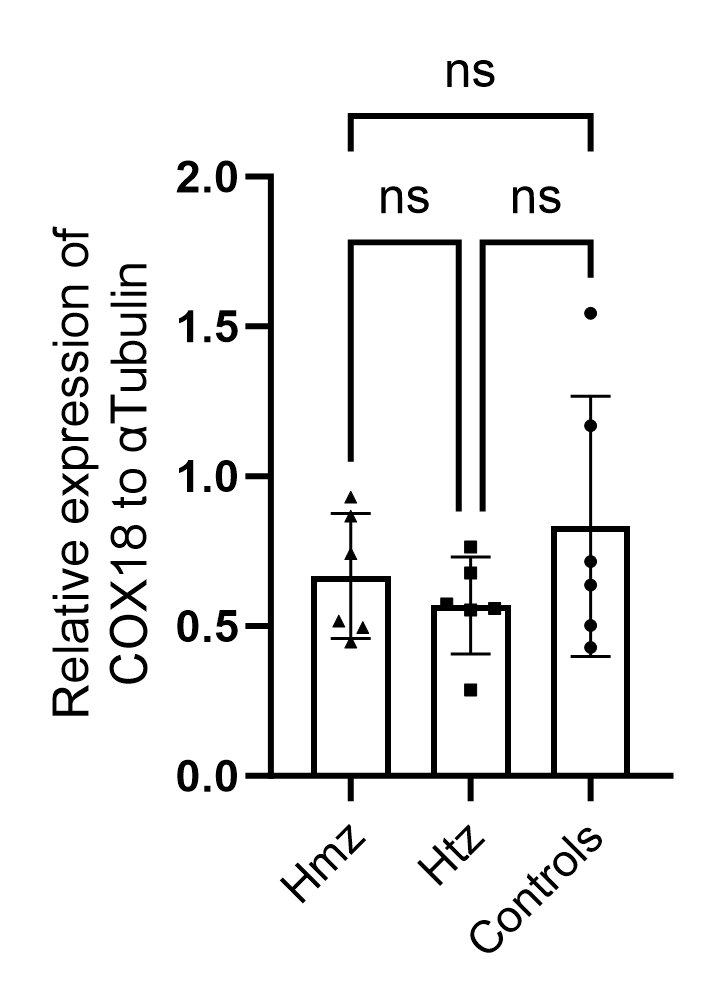

**Supplementary Figure 2.** **Immunoblotting of full lysate protein from EBV-transformed lymphoblasts derived from the 3 genotypes shows no reduced levels of COX18.** Quantification of the relative expression of Cox18 to alfa-Tubulin levels from full protein lysates. Data are shown as mean ± SD (n = 3 for each genotype, with 2 biological replicates for each genotype). ns = not significant.
